# Sub-Regional Corpus Callosum Morphology in Marijuana Users

**DOI:** 10.1101/2025.03.10.25323638

**Authors:** Mayan Bedggood, Florian Kurth, Eileen Luders, Mangor Pedersen

## Abstract

**Introduction:** The primary psychoactive component in marijuana is tetrahydrocannabinol (THC), which acts on receptors, such as cannabinoid 1 (CB1), that are distributed broadly throughout the brain. THC interferes with synaptic plasticity and neurogenesis and impacts the brain’s macrostructure, specifically white matter where CB1 receptors are abundant. The current study aims to investigate whether callosal morphology differs depending on how much experience individuals have with marijuana.

**Methods:** This is a quantitative between-group corpus callosum morphology analysis using cohort study data. The data for this study (*n* = 144) came from the S1200 Release from the Washington University – University of Minnesota Human Connectome Project Consortium (WU–Minn HCP). Marijuana use was quantified using self-reports and grouped as (1) no use, (2) low use, and (3) high use. T1-weighted MRI brain images were obtained and then processed using SPM12 and MATLAB. Each corpus callosum was manually traced and automatically separated into seven callosal areas according to the Witelson parcellation scheme. The resulting area measures were compared between the three groups, while covarying for total brain volume.

**Results:** Our ANCOVA analysis was significant (F(2, 145) = 4.38, *p* = .014), and posthoc tests revealed a significantly smaller anterior midbody in the high marijuana use group compared to the no marijuana use group (*p* = .012).

**Conclusion:** Smaller callosal areas in the high use marijuana group suggest that heavy cannabis may be related to weaker interhemispheric connectivity. Future research is required to replicate the current findings using well-powered designs.

## INTRODUCTION

Marijuana is the most widely used illicit substance globally (Brumback et al., 2016). The primary psychoactive component in marijuana is tetrahydrocannabinol (THC), which affects the endogenous cannabinoid system via receptors such as cannabinoid 1 (CB1), distributed broadly throughout the brain. THC can interfere with synaptic plasticity and neurogenesis, impacting brain volume and altering neural network maturation (Brumback et al., 2016). In particular, major targets of THC-mediated neurodegeneration seem to include white matter in the frontal lobes, fornix, hippocampus, frontal-limbic connections, corpus callosum, and commissural fibers, where CB1 receptors are abundant (Mandelbaum & de la Monte, 2017).

Previous studies have explored alterations in the corpus callosum, the brain’s most prominent white matter tract, with marijuana users. Orr et al. (2016) used Human Connectome Project data to look at marijuana use along a spectrum. The group comparison showed no significant differences and no linear effects of the number of times marijuana was used on white matter coherence in users. Orr et al. however, did not segment the corpus callosum into subregions and only looked at the whole structure. Gruber et al. (2014) also investigated white matter microstructure with chronic, heavy marijuana use. They showed that marijuana users had reduced fractional anisotropy, a measure of white matter fiber coherence (Gruber et al., 2014), compared to the control group in several regions, including the genu of the corpus callosum. In addition, Arnone et al. (2008) looked at mean diffusivity (MD), a measure of the average mobility of MD diffusivity increased in marijuana users in a corpus callosum region proximal to where white matter pathways connect the frontal lobes. Increased MD is usually consistent with increased water content, generally due to neuroinflammation or axonal loss (Arnone et al., 2008; Rigucci et al., 2016). Therefore, increased MD could indicate slower, less efficient information transfer (Rigucci et al., 2016).

Rigucci et al. (2016) examined the impact of cannabis on corpus callosum microstructural organisation in those with first-episode psychosis and those without. In addition to MD, they also looked at axial diffusivity (AD), which relates to the diffusion of water molecules parallel to the white matter fiber tracts. AD can be related to axonal function and whether the underlying bundles of white matter fibers are healthy (Seo et al., 2019). Rigucci et al.’s results showed that high-potency cannabis users had higher total corpus callosum MD and AD than low-potency users and those never used.

Increased AD could be due to increased extracellular water content as a result of white matter fiber atrophy and axonal loss (Seo et al., 2019). Subsequently, Rigucci et al.’s results showed that cannabis use was associated with a disturbed callosal microstructural organisation.

Lastly, Solowij et al. (2011) examined the effects of long term, heavy cannabis use on cerebellar structure in patients with schizophrenia and also in healthy individuals. They recruited 17 patients with schizophrenia and 31 controls, with 48% of the controls and 47% of the patients being long-term, heavy cannabis users. Using structural 3T MRI, they demonstrated reduced cerebellar white-matter volume in the cannabis users with and without schizophrenia compared to healthy controls (by 29.7% and 23.9% respectively) (Solowij et al., 2011).

While there is existing research looking into the possible impacts of marijuana use on white matter structures such as the corpus callosum, there are notable limitations of existing studies that warrant additional research. Many previous studies have used small sample sizes and often only included male participants (e.g., Arnone et al., 2008; DeLisi et al., 2006). This reduces the external validity of their results and means that the findings cannot be confidently applied to broader populations. In addition, many studies did not look specifically at the corpus callosum, or did not segment the corpus callosum into subregions (e.g., DeLisi et al., 2006; Gruber et al., 2014; Orr et al., 2016; Rigucci et al., 2016). Furthermore, some studies specifically looked at heavy marijuana use, without much focus on low usage (e.g., Arnone et al., 2008; Gruber et al., 2014).

Therefore, further research into the possible impact of marijuana on the different regions of the corpus callosum is required. Lastly, most of the evidence of a potential link between cannabis use and white matter damage comes from research using diffusion tensor imaging (DTI) to look at microstructural changes. More research is required to determine if there is a link between cannabis use and gross structural changes in white matter regions such as the corpus callosum. For the current study, we look at gross anatomical changes and hypothesise that subregions of the corpus callosum will be smaller for marijuana users compared to non-users, with more prominent effects in the high-use group.

## METHODS

### Data

The study sample was selected from the S1200 Release from the Washington University – University of Minnesota Human Connectome Project Consortium (WU– Minn HCP) (Van Essen et al., 2013). This sample was taken from a healthy population, and an overview of the participant recruitment strategy is detailed elsewhere (Van Essen et al., 2013). Ethics for the project were approved by the host institutions (Van Essen et al., 2013). As the present study addresses non-clinical, non-sensitive and non-identifiable data, no further ethics were required beyond what was already obtained for the original data collection. The variable of interest was the use of marijuana, which was quantified using self-reports as assessed by the Semi-Structured Assessment for the Genetics of Alcoholism (SSAGA) (Bucholz et al., 1994). This tool recorded participants’ marijuana use on an ordinal scale ranging from 0 to 5 (never used = 0; used 1 to 5 times = 1; used 6 to 10 times = 2; used 11 to 25 times = 3; used 26 to 50 times = 3; used 51 to 100 times = 3; used 101 to 999 times = 4; used 1000 to 2000 times = 5 and used more than 2000 times = 5). For the current study, we categorised marijuana use into three groups. For each participant, their level of marijuana use was classified as (1) no use, (2) low use (if they had reported using marijuana between 1 and 100 times total in their lifetime), and (3) high use (if they had reported using marijuana more than 100 times total in their lifetime). The high use group was the smallest, with 48 subjects (*mean age* = 28.3, *SD* = 3.99), therefore, we randomly sampled 48 participants from the no use group (*mean age* = 28.3, *SD* = 3.96) as well as the low use group (*mean age* = 28.3, *SD* = 3.96) to compile three groups that are matched for age and sex (24 males, 24 females each). This resulted in a total sample size of 144 for the group comparisons.

### Image Acquisition and Pre-Processing

High-resolution T1-weighted brain images (0.7×0.7×0.7 mm^3^) were obtained on a customized Siemens 3T Connectome Skyra scanner at Washington University with a 32-channel head coil (TR = 2400 ms; TE = 2.14 ms; FOV = 224 × 224). All brain images were processed using SPM12 and MATLAB R2022B, applying bias field corrections and rigid-body spatial normalisations.

### Callosal Tracing

Each corpus callosum was manually traced at the mid-sagittal brain section by a researcher (M.B.) blinded to the participant’s group status. This began with a training phase, where 20 individual corpus callosum were traced and both intra- and inter-rater variability were assessed (with index scores of 0.966 and 0.962, respectively). Once the real dataset had been traced, the Witelson parcellation scheme was then applied (see Figure 1 and Witelson et al., 1989), which resulted in seven callosal segments (rostrum, genu, rostral body, anterior midbody, posterior midbody, isthmus, and splenium). We then calculated the areas (in mm^2^) for each callosal segment, in addition to the total area of the corpus callosum (total CC).

**Figure 1.**
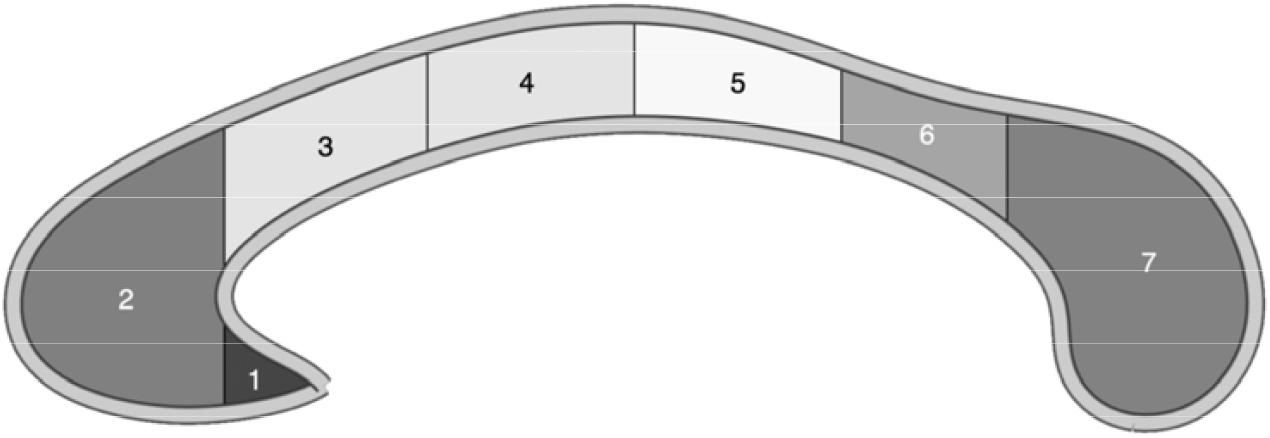
Callosal outline and segmentation based on the mean callosal outline of all participants *Note:* The corpus callosum was separated into seven regions using the Witelson parcellation scheme: (1) rostrum, (2) genu, (3) rostral body, (4) anterior midbody, (5) posterior midbody, (6) isthmus, and (7) splenium.

## Statistical Analysis

An analysis of covariance (ANCOVA) was conducted for each callosal subregion and the total corpus callosum to determine if there were any significant differences in callosal area size between the three marijuana groups. For each of the ANCOVAs, the independent variable was marijuana use with three levels (no use, low use, and high use), the covariate was total intracranial volume (TIV), and the dependent variable was the subarea measurement (i.e., rostrum, genu, rostral body, anterior midbody, posterior midbody, isthmus or splenium) or total area measurement. Follow up post hoc tests were conducted for any significant omnibus ANCOVA result using Jamovi Version 2.3.19.0.

## RESULTS

The only significant ANCOVA result was a difference in the area of the anterior midbody across the three marijuana groups (no use, low use and high use), F(2, 145) = 4.38, *p* = .014. Follow-up tests revealed a significantly smaller anterior midbody (*p* = .012) in the high marijuana use group (*M* = 83.8, *SD* = 12.5) compared to the no marijuana use group (*M* = 91.6, *SD* = 15.3), as shown in Figure 2.

**Figure 2.**
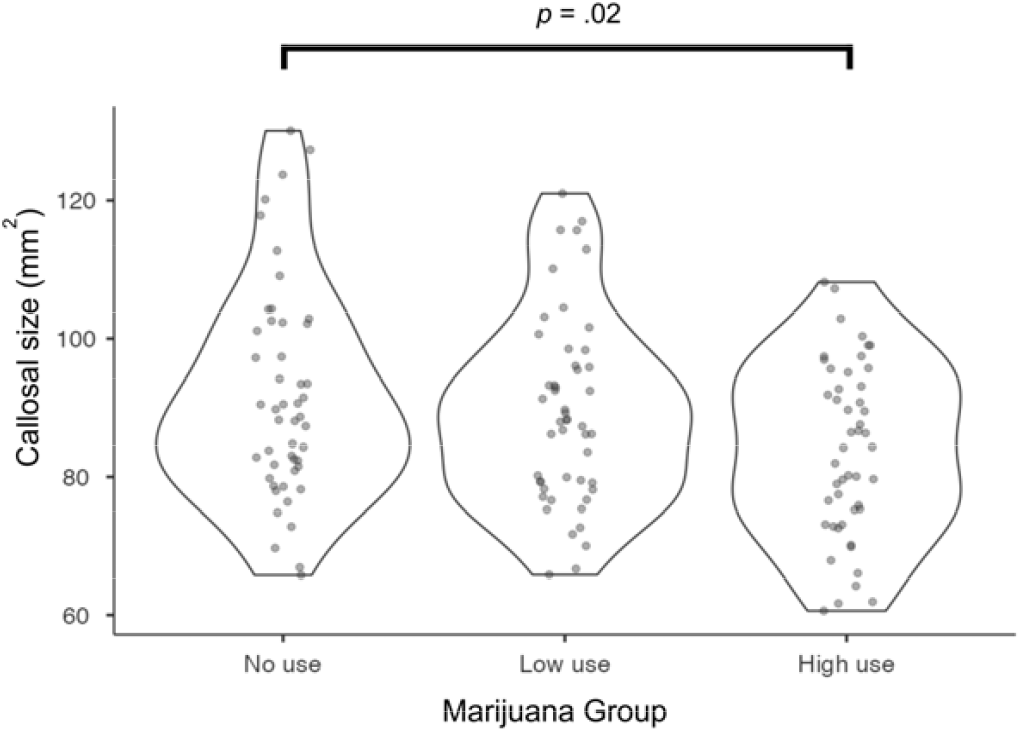
Smaller Callosal Anterior Midbody in High Marijuana Users *Note*. Belween-group comparison (no marijuana use; low marijuana use; high marijuana use) of sub-regional callosal morphology in the anterior midbody region. The violin plot reflects the group-wise data distribution and each dot denotes a single subject.

## DISCUSSION

Our results indicate that the anterior midbody may be smaller for the group with high marijuana use than the group that had never used marijuana. The corpus callosum has a functional topographic organisation (Fabri et al., 2011; Mara, & Gabriele, 2023), and current research indicates that premotor and supplementary motor areas, and therefore motor function, may be reliant on fibers that cross through the anterior midbody (Fabri et al., 2011; Mara, & Gabriele, 2023). A smaller anterior midbody may therefore suggest weakened interhemispheric communication between motor areas.

Our results align with research that links heavy, frequent cannabis use and disturbed callosal microstructural integrity (Rigucci et al., 2016). For example, Rigucci et al. (2016) found differences in the posterior part of the corpus callosum, in the splenium and posterior midbody. Zalesky et al. (2012) also found changes in the splenium of the corpus callosum in people with a long history of heavy cannabis use. Furthermore, Arnone et al. (2008) found microstructural changes in marijuana users compared with controls, specifically in the callosal fibers that connect the prefrontal lobes. These findings support the suggestion that marijuana may be linked to impaired structural integrity. However, they differ from the current findings in terms of the exact location of the changes.

Regarding the neurobiological etiology, there is significant focus in the current literature on the CB1 receptors that THC innervates. These receptors are prolific in white matter and have known effects on oligodendrocyte development, which impacts the myelin surrounding axons (Rigucci et al., 2016). Mechanistically, endocannabinoid receptors such as CB1 and CB2 modulate synaptic plasticity and cortical connectivity (Mandelbaum & de la Monte, 2017). The THC in marijuana activates cannabinoid receptors in the brain (especially CB1), therefore interfering with the physiological actions of endocannabinoids (Mandelbaum & de la Monte, 2017; Testai et al., 2022).

This can disrupt the fine-tuning of endocannabinoid synaptic activity and impair the connectivity of neural networks (Testai et al., 2022). In terms of the difference between low and high marijuana use, chronic and/or heavy exposure to marijuana may result in the process of neuroadaptation (Arnone et al., 2008), whereby prolonged exposure to THC induces CB1 receptor downregulation and desensitization (Testai et al., 2022).

Furthermore, long-term exposure to THC facilitates microglia and astrocytes to release inflammatory cytokines and to reduce the activity of anti-inflammatory cytokines (Testai et al., 2022). Therefore, the neurobiological etiology of the marijuana-linked alterations in callosal white matter could be a combination of increased CB1 receptor activation, subsequent downregulation, and increased extracellular water content due to neuroinflammation and edema. Both processes could result in axonal atrophy, indicated by increased diffusion parameters in previous research (Arnone et al., 2008; Gruber et al., 2014; Rigucci et al., 2016; Seo et al., 2019).

Lastly, research has supported that marijuana, specifically, may be responsible for these neural changes. For example, Rigucci et al. (2016) demonstrated that high cannabis use was associated with a disturbed callosal microstructural organisation. Importantly, they found that this result was not dependent on whether participants were in the psychosis group, suggesting that the impact on the corpus callosum was not due to the psychosis. In addition, Becker et al. (2015) found that heavy cannabis use was associated with disturbances in areas such as the corpus callosum. Notably, although the group that used cannabis also consumed more alcohol than the control group, it was found that only their cannabis use was quantitatively associated with microstructural change. These results strengthen the inference that cannabis use could be a primary factor underlying potential changes in white matter.

Strengths of the current study include that our study looked at high and low marijuana use instead of simplifying it to marijuana use versus no use. This enabled the recent research to delineate further whether any significant findings were with specific frequencies of marijuana use or not. For example, the fact that there was no significant difference in callosal morphology between low marijuana use and no use could indicate that the harmful effects of marijuana use are exacerbated by higher or more prolonged use. Limitations of the current study include the narrow age range (22 – 36 years) and the substantial difference in marijuana use frequency between the groups. Future research could investigate corpus callosum morphology in the broader age group and conduct longitudinal follow-ups. This could help determine whether any observed anatomical variations normalise if marijuana use is ceased or, conversely, if they seem irreversible.

## CONCLUSION

The current study’s findings indicate that the corpus callosum’s anterior midbody region may be smaller in those with high marijuana use. This may indicate that the interhemispheric communication facilitated by the fibers from that region, such as premotor and supplementary motor areas, are weakened in those with high marijuana use. The neurobiological basis could be downregulation and increased extracellular water content due to neuroinflammation, with subsequent axonal loss. Future research could replicate these findings and investigate this effect in a longitudinal study to see if any detrimental impacts are reversible with cessation of marijuana use.

## Data Availability

All data produced are available online at https://www.humanconnectome.org/

https://www.humanconnectome.org/

## Acknowledgments

Data were provided by the Human Connectome Project, WU-Minn Consortium (Principal Investigators: David Van Essen and Kamil Ugurbil; 1U54MH091657) funded by the 16 NIH Institutes and Centers that support the NIH Blueprint for Neuroscience Research; and by the McDonnell Center for Systems Neuroscience at Washington University.

